# Lifestyle Empowerment for Alzheimer’s Prevention Prescribed by Physicians: Methods and Adaptations to COVID-19

**DOI:** 10.1101/2024.07.29.24311181

**Authors:** Amanda N. Szabo-Reed, Amber Watts, Eric D. Vidoni, Jonathan Mahnken, Angela Van Sciver, Katrina Finley, Jonathan Clutton, Rachel Holden, Mickeal N. Key, Jeffery M. Burns

## Abstract

The health care system is insufficiently capitalizing on the benefits of physical exercise in America’s aging population. Few tools exist to help clinicians incorporate physical activity into their clinical care, while barriers limit older adults from initiating and maintaining exercise programs. The Lifestyle Empowerment for Alzheimer’s Prevention (LEAP! Rx) Program has been designed to support providers and participants in lifestyle change. LEAP! Rx uses two forms of participant enrollment: physician referrals through electronic health records and self-referrals to test the efficacy of delivering a community-based exercise and healthy lifestyle program to older adults. After referral into the program, participants are randomized to receive the LEAP! Rx Program or are placed in a standard-of-care group to receive the program later. The LEAP! Rx program consists of a personalized and structured exercise program, lifestyle education, and mobile health monitoring. This includes a 12-week Empowerment phase with coaching and supervised exercise training, followed by a 40-week Lifestyle phase with intermittent supervised exercise and coaching. Lifestyle education includes monthly, evidence-based classes on optimal aging. The evaluation of LEAP! Rx focuses on 1) the assessment of implementation and scalability of the LEAP!Rx Program for clinicians and patients 2) the effect of the LEAP! Rx Program on cardiorespiratory fitness, 3) the impact of the LEAP! Rx Program on secondary intervention outcome measures of chronic disease risk factors, including insulin resistance, body composition, and lipids. If successful, this study’s findings could advance future healthcare practices, providing a new and practical approach to aging and chronic disease prevention.

## 1.0 Introduction

The unprecedented growth of the elderly population underscores the need for innovation in promoting health in America’s aging population.[1, 2] Americans live longer than ever. [3] Approximately 20 percent of the population is 65 years and older, and this age group will continue to grow.[4] Aging brings increased cognitive and physical decline with dramatic rises in the risk of cardiovascular disease, stroke, diabetes, and Alzheimer’s disease (AD). [1] There is strong evidence that lifestyle interventions can modify these risks. [5]

### 1.1 Lifestyle Interventions Represent Promising Prevention Strategies

Ongoing research suggests that modifying lifestyle behaviors represents an essential strategy for preventing age-related chronic disease. [5] Physical activity is among the most important and cost-effective tools available for chronic disease management.[6] with its well-described effects on diabetes,[7, 8] obesity,[9–12] hypertension,[13] and depression.[14, 15] Recent research suggests exercise also benefits the brain.[16–19] Increased cardiorespiratory fitness in early Alzheimer’s disease (AD) is associated with increased whole-brain volume,[16] most strongly in regions relevant to AD (the hippocampus and parietal lobe).[17] Longitudinal studies suggest a decline in cardiorespiratory fitness is associated with brain atrophy in early AD and cognitively normal individuals.[18] Insulin levels are associated with cognitive performance and brain volume.[20–22] Specifically, physical activity modulates several important vascular risk factors – atherosclerosis,[23] heart disease,[24] stroke,[25] diabetes[26–31], inflammation,[32] and insulin signaling[33–36] – that represent essential components of age-related disease.[37]

### 1.2 Public Health Efforts Currently Fall Short

There has been little systematic progress in translating lifestyle intervention research into meaningful gains in population health. While lab-based trials demonstrate efficacy in highly controlled environments, they are often impractical when translated to the broader population. Many clinicians recommend increased physical activity to their patients. This recommendation, however, lacks specificity[38], and numerous barriers conspire against older adults to limit physical activity.[39] In addition, recommendations from clinicians are often singular events with little follow-up. The current approach of education and encouragement alone is insufficient: up to 25% of all adults in the U.S. are entirely sedentary, and more than half are insufficiently active to achieve health benefits [6, 40] with physical activity levels declining through adulthood to low levels in older adults,[6, 41, 42] exacerbating the risk of functional decline.[43] Therefore, it is essential we provide clinicians with specific, practical tools to help their patients initiate and sustain healthy lifestyle behaviors. This requires innovation in optimizing the delivery of lifestyle-based interventions to the public. A critical gap exists for patients to navigate between the clinician’s office and the resources to support a healthier lifestyle (i.e., fitness centers, trainers, dietitians). We have designed the LEAP! Rx Program to fill that gap, building on documented success when clinicians specifically prescribe exercise.[44–47]

### 1.3 Importance of an Individualized, Community-Based Approach

LEAP! Rx is a flexible approach using community-based fitness resources to help individuals overcome barriers such as motivation, access, and support. The LEAP! Rx Program leverages the patient’s clinician, familiar institutions, a supervised exercise prescription, and monitoring as entry points to initiate and sustain habitual physical activity. The program relies on three design features to encourage sustained changes in health behavior: individualization, feedback, and accessibility. Personal LEAP! Rx Coaches trained in motivational interviewing assess readiness for change[48] and identify individual motivations, goals, and barriers to sustained physical activity. Feedback is essential in supporting the intervention through self-monitoring physical activity with feedback loops to LEAP! Rx Coaches and clinicians. Accessibility is optimized through community-based fitness centers that provide ready expertise and infrastructure at low cost in a community-based setting close to home.

### 1.4 Summary

Currently, the healthcare system is insufficiently capitalizing on the potential of lifestyle changes to reduce the population burden of chronic disease in older adults. Simply encouraging older adults to adopt healthier lifestyles is not enough. Innovation is needed to develop interventions to maximize successful aging through strategies to sustain older adults’ health and function.[49] Hence, there is a critical need to translate the latest evidence-based approaches into broad public health benefits. The primary purpose of this study is to test a framework for clinicians to engage their patients in healthy behaviors and sustained lifestyle changes following public health recommendations (see Figure 1). We anticipate the framework will improve important health outcomes in older adults, with benefits on multiple levels:

1. To healthcare providers who need practical tools to promote prevention strategies in older adults;
2. To patients who need guidance on evidence-based strategies to maintain optimal health through proven programs that help initiate and sustain behavior changes;
3. To healthcare systems needing proven options for preventative health programs and guidance on leveraging them (i.e., referral process, links to community-based programs).

**Figure 1.**
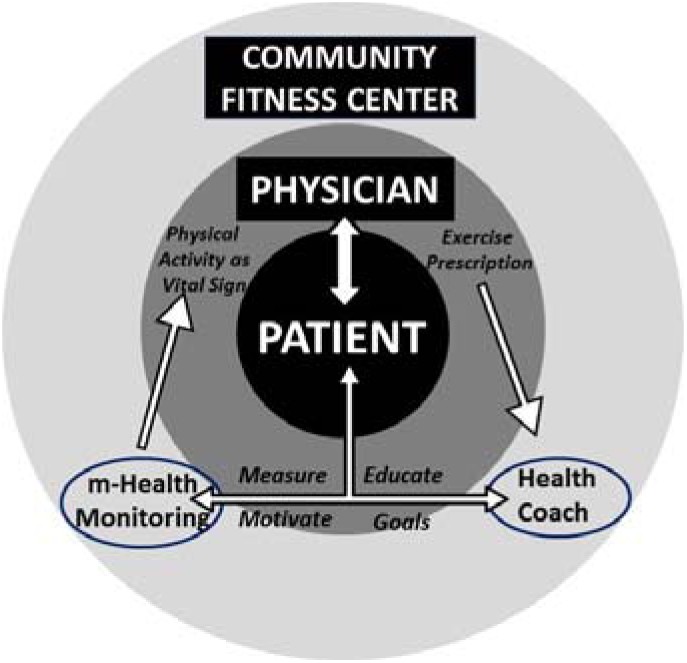
The LEAP! Rx Program supports the clinician-patient relationship. The clinician prescribes the program of coaching, training, and monitoring in a community based fitness center that feeds easily-digested metrics back to the clinician to encourage long term compliance.

Additionally, if this approach reduces the burden of chronic disease, it could have significant public health and economic consequences. Based on emerging data in the field of cognitive aging, 40% of AD cases are attributable to modifiable risk factors,[50] and even a modest 10 – 25% reduction in these risk factors is estimated to cut the number of adults in the US with AD by nearly 500,000 cases.[51] Thus, optimizing lifestyle strategies to maximize their effect will likely translate into sizeable public health gains.

### 1.5 Hypotheses

The goals of this study are to:

1. To asses the implementation and scalability of the LEAP!Rx Program for clinicians and patients. We will test the efficacy of an exercise and healthy lifestyle program using a unique referral method embedded in the electronic health record (EHR) as compared to the standard of care.
2. Determine the effect of the LEAP! Rx Program on cardiorespiratory fitness. We hypothesize that the LEAP! Rx Program will increase and maintain cardiorespiratory fitness (peak oxygen consumption [VO_2peak_]) at both 12 weeks (after the initiation phase) and 52 weeks (after the maintenance phase).
3. Test the effect of the LEAP! Rx Program on outcome measures of chronic disease risk factors, including insulin resistance, body composition, and lipids. We hypothesize that LEAP! Rx Program will improve an individual’s metabolic profile with measurable benefits in specific secondary outcome measures of HOMA2, fat mass, lean mass, and cholesterol (total, LDL, HDL).

## 2.0 Methods

Our goal is to randomize two hundred twenty participants to the LEAP! Rx Program vs. a standard of care control group (1:1 ratio, Figure 2). The standard of care group receives educational materials at entry – mimicking the current standard of care for encouraging health behavior change – and provide outcome measures parallel to those of the active intervention group. After completion of the study, this group has access to the LEAP! Rx Program (to enhance recruitment efforts; no outcomes measured).

**Figure 2.**
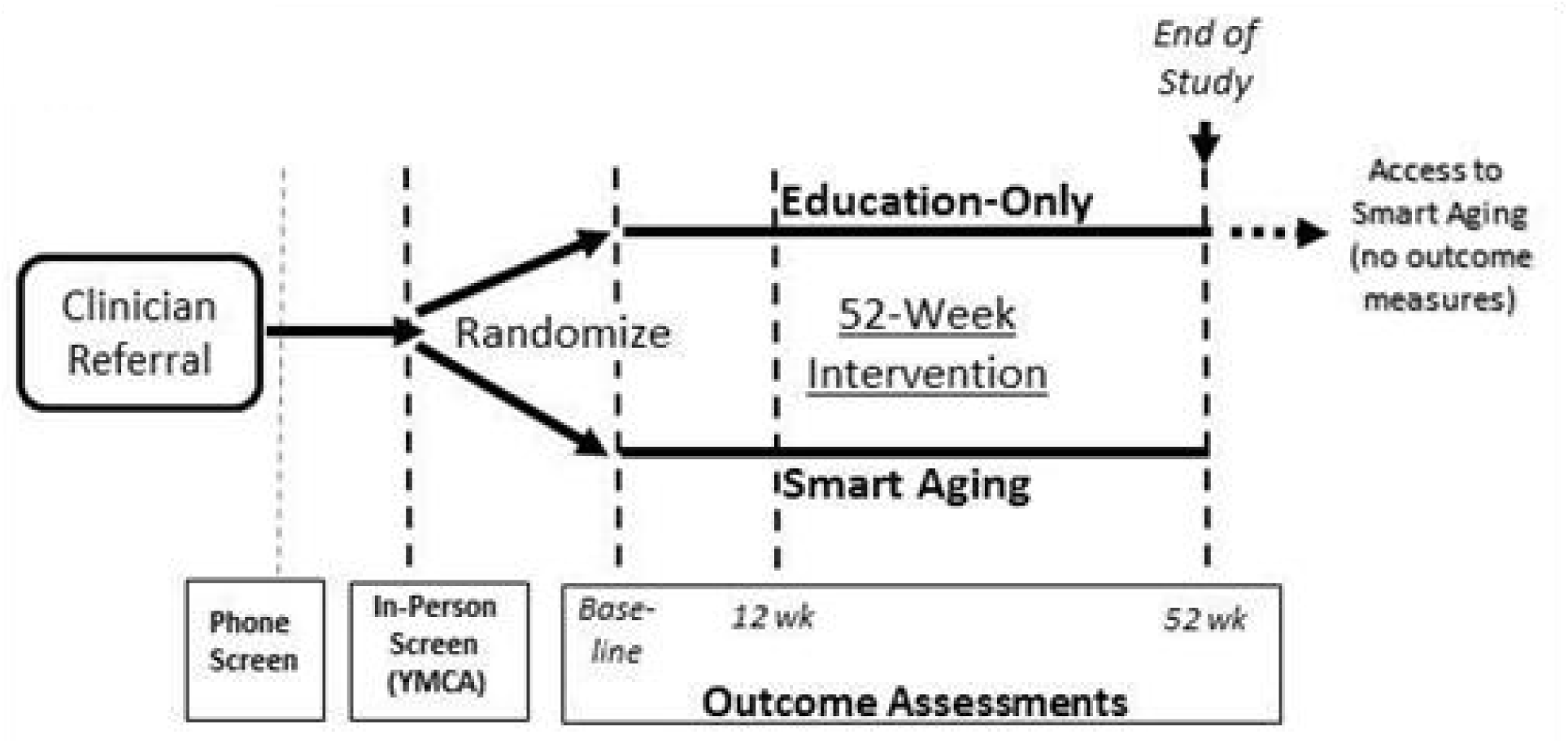
Study flow.

### 2.1 Inclusion/Exclusion

The inclusion/exclusion criteria (Table 1) are intentionally broad to provide a generalizable cohort of older adults and encourage clinician referral of participants who the clinician believes would benefit from increased physical activity. Participants are clinician-referred or self-referred, ambulatory, underactive, and 65 years and older without a recent history of stroke or coronary artery disease.

**Table 1.**
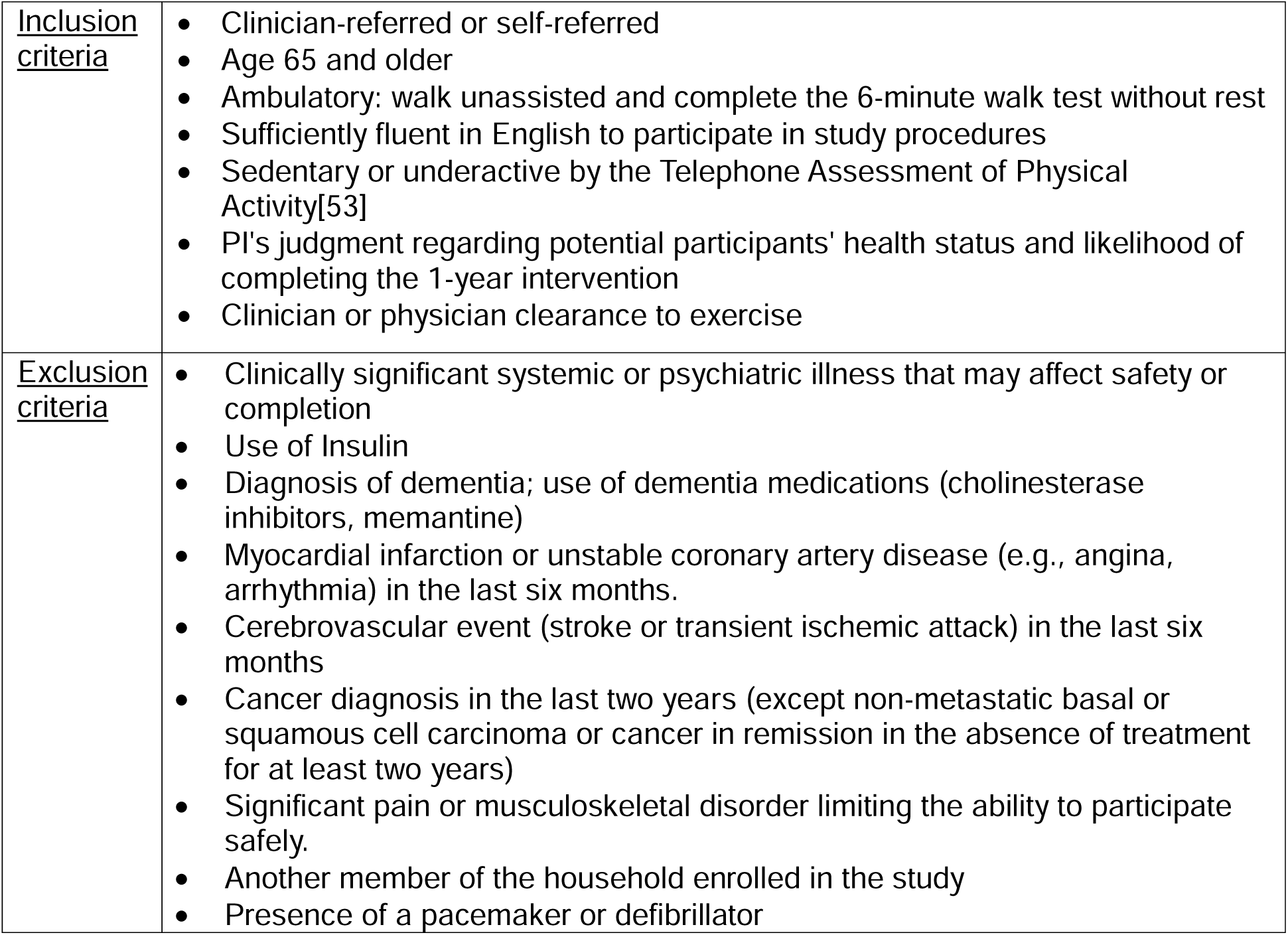
Inclusion/Exclusion Criteria.

|  |  |
| --- | --- |
| <u>Inclusion criteria</u> | • Clinician-referred or self-referred<br>• Age 65 and older<br>• Ambulatory: walk unassisted and complete the 6-minute walk test without rest<br>• Sufficiently fluent in English to participate in study procedures<br>• Sedentary or underactive by the Telephone Assessment of Physical Activity[53]<br>• PI's judgment regarding potential participants' health status and likelihood of completing the 1-year intervention<br>• Clinician or physician clearance to exercise |
| <u>Exclusion criteria</u> | • Clinically significant systemic or psychiatric illness that may affect safety or completion<br>• Use of Insulin<br>• Diagnosis of dementia; use of dementia medications (cholinesterase inhibitors, memantine)<br>• Myocardial infarction or unstable coronary artery disease (e.g., angina, arrhythmia) in the last six months.<br>• Cerebrovascular event (stroke or transient ischemic attack) in the last six months<br>• Cancer diagnosis in the last two years (except non-metastatic basal or squamous cell carcinoma or cancer in remission in the absence of treatment for at least two years)<br>• Significant pain or musculoskeletal disorder limiting the ability to participate safely.<br>• Another member of the household enrolled in the study<br>• Presence of a pacemaker or defibrillator |

### 2.2 Referral and Recruitment Process

#### 2.2.1 Referral Process

We recruit participants by two modes: direct referral from clinicians (i.e., physicians, nurse practitioners) of the hospital or the general population via typical recruitment methods such as advertisement, flyers, and in-person talks. Our referral process for clinicians is embedded in the EHR as a “prescription” so that the LEAP! Rx Program is ordered as part of a seamless clinical workflow. Workflow development included 1) identifying precise, computable phenotypes to identify potential participants, 2) flagging these potentially eligible participants for providers (“best practice alert”), and 3) routing the prescription from the health system through the study team with minimal delay, and on to activation for enrollment at the YMCA. The ideal flow would be direct from provider to community support. However, for research rigor, the electronic prescription or recruitment division referral to the study team allows for initial contact to determine eligibility and outcome test scheduling. A clinician or physician clearance to exercise is required for self-referred participants.

#### 2.2.2 Clinician Role

Clinicians “prescribe” the program to potentially eligible patients via an electronic prescription, just like other orders in the EHR. Evidence suggests that a brief 3-4 minute office discussion is meaningful without adding a significant burden and can aid in program recruitment and adoption.[52]

#### 2.2.3 Phone Screening

KU study staff contact participation candidates to assess eligibility, obtain permission to examine their health records, and perform a Telephone Assessment of Physical Activity [53] to ensure participants are sedentary or underactive.

#### 2.2.4 Enrollment and Randomization

After completing the phone screen, eligible participants are given a standard set of informational materials on the study. If the potential participant is interested in participating in the study, study staff schedule the baseline visit. Baseline testing is scheduled with a goal of completion within a two-week window. Upon completing baseline testing, the study coordinator randomizes participants to either the active intervention or the standard of care 1-year wait-list control group (1:1 ratio). The randomization table with random block sizes was developed in SAS by the study statistician and uploaded to the data capture system, REDCap, to allocate participants to the treatment arm securely.

#### 2.2.5 Blinding

Psychometrists and exercise physiologists who perform assessments are blinded to intervention groups. The primary investigator is unblinded to perform safety assessments and address safety concerns or adverse events but is not involved in acquiring outcome measures and does not have access to the outcome measures in REDCap.

### 2.3 Control Group: Standard of Care

The standard of care group has been designed to represent the current standard of clinical care. Individuals randomized to this group are only given educational information on the benefits of a healthy lifestyle at baseline, but are not discouraged from exercising independently. However, we do ask that they agree not to join a YMCA or hire a personal trainer during this time. Current data suggest that education only does not result in significant and sustained physical activity changes.[44, 54, 55]

Participants randomized to the standard-of-care group are informed they have been randomized to a 52-week “waiting list,” during which time they complete all outcome measures parallel to the active intervention group. All participants (both active and standard of care) receive the same educational materials at the enrollment visit. After completing outcome assessments at 52 weeks, the standard-of-care participants are offered access to the full LEAP! Rx Program at no cost, including membership and coaching fees, to enhance recruitment and minimize dropout from this group. Outcome assessments are not repeated during their participation in the LEAP! Rx Program.

Cross-over does not threaten internal validity as the study is designed to assess the value of the LEAP! Rx Program over and above the effects of standard of care (i.e., education only). Control participants can exercise independently (though no structured assistance is provided for this comparison group).

### 2.4 Intervention Group: LEAP! Rx Program

The LEAP! Rx Program is packaged to be delivered at any fitness center. For this study, partner with local YMCA sites.The LEAP! Rx Program represents a “package” consisting of:

1. A dedicated LEAP! Rx Coach (i.e., personal trainer)
2. Mobile health objective monitoring of daily physical activity
3. Smart Aging Educational Curriculum
4. Group exercise opportunities

Additional details about the LEAP!Rx program can be found in Supplemental Material 1.

### 2.5 Response to the COVID-19 Pandemic

In March 2020, exercise facilities were closed in response to the COVID-19 pandemic. Many facilities remained closed until the end of May 2020. During this time, participants continued meeting with their LEAP! Rx Coach virtually over Zoom to complete exercise sessions at home. In addition, in-person education curriculum sessions were discontinued and only offered online. After May 2020, those participants who were comfortable and interested in resuming in-person exercise did.

### 2.6 Study Events

#### 2.6.1 Overview

Physiologic adaptations to the LEAP! Rx Program are assessed with cardiorespiratory fitness testing, physical function assessment, body composition measures, laboratory tests, health surveys, and cognitive testing (Table 2). Study assessments are completed in one appointment visit lasting approximately two hours.

#### 2.6.2 Baseline, Week 12, and Week 52 Outcome Assessments

Baseline, 12-week, and 52-week outcome assessments are identical and included in Table 2. Briefly, consented participants had fasting phlebotomy and were provided with a snack. Trained research staff collect demographic information, health history, and medications at baseline (and update at follow-up visits). Blood pressure (measured three times in a sitting position), height, and weight are assessed each time. Trained staff complete DEXA scanning and cardiorespiratory fitness testing with graded maximal exercise testing. Participants perform less than 2 hours of surveys (NIH Toolbox Emotion, diet survey, skin carotenoid spectroscopy (Veggie Meter®, Longevity Link Corp), and brief cognitive evaluation (NIH Toolbox Cognitive). These procedures take approximately two hours to complete in a single visit. An accelerometer is placed on the participant at the first visit and removed at the second visit unless extenuating circumstances arise. In this case, the accelerometer is mailed with wear instructions.

#### 2.6.3 Cardiorespiratory Fitness

Our primary intervention outcome is cardiorespiratory fitness as measured by peak oxygen consumption (VO_2 peak_) during treadmill testing at 12 and 52 weeks compared to baseline. This co-primary outcome will test our hypothesis that the LEAP! Rx Program will increase VO_2 peak_ after the intensive Empowerment phase (12 weeks) and remain increased over baseline during the Lifestyle phase (52 weeks).

VO_2 peak_ is measured during the graded treadmill test to volitional exhaustion or following American College of Sports Medicine (ACSM) guidelines [56]. The test is led by an exercise physiologist and supervised by a clinician using a modified Bruce protocol.[57] Participants are attached to a 12-lead electrocardiograph and wear a non-rebreathing facemask to assess heart rate, blood pressure, and expired air (TrueOne, Parvomedics, Sandy UT). VO_2 peak_ is the highest observed value of oxygen utilization during the test.[58, 59]

#### 2.6.4 Physical Activity Measures

##### Accelerometry

We use wrist-worn triaxial accelerometers (Actigraph GT9X, Pensacola, FL) during three 1-week periods (baseline, week 12, and week 52). We have chosen wrist-worn devices (as opposed to hip or thigh devices) based on our experience with both wrist and hip-worn devices[60] and recent literature relevant to older adults.[61] Wrist-worn accelerometers are comparable to hip-worn when assessing moderate-to-vigorous physical activity (MVPA) AND are associated with better adherence.[61] Triaxial accelerometers are more thoroughly validated to provide a research-based outcome measure than consumer products (such as pedometers used for self- and clinician monitoring). Our accelerometer protocol was adapted from the NHANES study[62] and we have successfully collected data from over 100 early AD patients and cognitively normal older adults.[60]

The accelerometer are worn on the non-dominant wrist during a consecutive 7-day period. Participants are instructed to wear the unit 24 hours a day. The unit is waterproof and can be worn while bathing and swimming. Total daily activity (sum of all activity during waking hours), the intensity of daily activity (divide total activity by the time of all nonzero epochs), and the percent of the day spent in non-activity will be calculated using ActiLife software[63] including the selection of proper epochs (1 minute[64]), length of activity bouts, intensity thresholds, data transformation, and missing data imputation.[65] We consider days with > 10 hours of wear time to be a valid day and require at least four valid days (including one weekend day) of observation.[66]

##### The International Physical Activity Questionnaire (IPAQ)

survey is administered to assess the participant’s habitual level of physical activity. IPAQ is a 27-item measure developed to assess physical activity levels in older adults. It is a robust measure (unaffected by seasonal bias) designed to capture various physical activities in older adults. This instrument is a valid and reliable method to assess changes in response to interventions.[67–69]

#### 2.6.5 Physical Function

After training by study staff, and demonstration of competence, the YMCA LEAP! Rx Coaches perform the 6-Minute Walk Test (6MWT), a well-known test of aerobic endurance[70] and has been shown to capture training responses.[71] The LEAP!Rx Coaches administer the 6MWT during the first intervention session at the YMCA and again at Weeks 12 and 52.

#### 2.6.6 Body Composition

Participants have their body composition measured using dual-energy X-ray absorptiometry (DEXA, Lunar Prodigy, version 11.2068, Madison, WI) to determine fat-free mass, fat mass, and percent body fat. Participants are weighed with a digital scale accurate to ±0.1 kg in a standard hospital gown after attempting to void and removing jewelry.

#### 2.6.7 Laboratory measures

A fasting blood draw (∼ 35mL total) is performed and processed to generate plasma and serum for biomarker analysis. Plasma and serum are stored at −80C for future analysis of glucose, insulin, lipids, and related metabolic markers.

#### 2.6.7 Patient-Centered Measures

Patient experience, psychosocial outcomes, attendance and dropout rates, satisfaction, quality of life, and self-efficacy are assessed at baseline and 12 and 52 weeks.

##### NIH Toolbox Emotion Battery

The NIH Toolbox Emotion Battery [72] includes a variety of measures that are used to assess participants’ responses to the LEAP! Rx Program. The battery consists of validated measures of General Life Satisfaction and Self-Efficacy. Additionally, it includes several relevant mental health measures such as Positive Affect, Sadness, Emotional Support, and Perceived Stress.

##### Health-related quality of life (HR-QOL)

refers to how health impacts an individual’s ability to function and perceived well-being in physical, mental and social domains. The Medical Outcomes Study 36-Item Short Form (SF-36) health survey is the most commonly used HR-QOL measure. The SF-36 health survey is a widely used health status questionnaire comprised of 36 items selected from a larger pool of items used by RAND in the Medical Outcomes Study (MOS).[73]

##### Dropout, Adherence, and Attendance

We monitor dropout rates. We anticipate measuring adherence as the number of exercise sessions completed (center-based training, group sessions, and home-based sessions), the number of scheduled exercise training sessions completed, and the number of LEAP!Rx Coaching sessions attended. Participants maintain paper-based logs to record their home-based exercise and group exercise attendance. LEAP!Rx Coaches enter data from the paper-based logs into a central database maintained by KU staff. Additionally, attendance at scheduled LEAP! Rx education classes (monthly), exercise training sessions, and LEAP!Rx Coaching sessions are recorded by YMCA staff and entered into the central database. Attendance data is shared with Referring Clinicians.

##### Exit Surveys

We also perform a short semi-structured exit survey on all patients who complete the study (at Week 52 visit) to assess their general experience, perceived benefits, and suggestions for improvement. Participants who drop out are interviewed by phone to ascertain their reasons for dropping out, perceived barriers, suggestions for program improvement, and general feedback.

#### 2.6.8 Clinician Measures

Because our overall object is to create a framework for clinical lifestyle prescription, we aim to create a clinician tool for the effortless “prescription” of the LEAP! Rx Program. In return, the clinician receives simple, easily digestible metrics on patient progress (attendance and physical activity data) to support the clinician-patient relationship.

Referral patterns are assessed per clinic type and clinician by examining the number of referrals, timing of referral, and clinical indication for referral. Examining the time and source of referrals will provide insight into adoption rates, whether program growth is organic vs. driven by our efforts to engage clinicians, and whether these adoption rates are variable across clinics.

#### 2.6.8 Other outcomes

##### NIH Toolbox Cognitive Function

Although not a primary aim of this project due to power issues, we will collect basic cognitive function data using a brief battery derived from the NIH toolbox. [74] This information may prove useful in powering a larger future study if needed.

##### Diet intake survey

As greater attention to one’s lifestyle may lead to changes in the composition of our participant’s diets, we assess changes in diet during the intervention using the NCI Diet History Questionnaire II as an exploratory outcome. Diet is strongly linked to diabetes, obesity, hypertension, and inflammation, while specific diets have been linked with AD and dementia outcomes. [75–78]

##### Skin Carotenoids

Skin carotenoids are measured at the baseline and final visits in a sub-sample of participants. [79] This tool uses reflection spectroscopy to measure skin carotenoid levels, which can be associated with fruit and vegetable consumption. The participant will place an index finger in the probe for 10 seconds while the skin tissue is measured. Three measurements are taken for an average score.

### 2.7 Data Management

Study data are collected and managed using web-based, electronic data capture tools hosted on a secure, HIPAA-compliant server. Data are collected on standard source documents or via direct data entry into REDCap hosted on a secure, institution server with role-based access.

### 2.8 Sample Size

Our overall goal is to create a framework for clinical lifestyle prescription. To demonstrate the effectiveness of the LEAP! Rx prescription and to create “buy in,” we seek to demonstrate the change in cardiorespiratory fitness. Thus, the primary analysis of our primary intervention measure – VO_2peak_ at both 12 and 52 weeks – is the basis for the sample size of our study. Our analysis will be an intent-to-treat comparison of active vs. control groups using a linear mixed model of repeated measures of VO_2peak_ at initial measurement, 12 weeks later, and after 52 weeks. Study power is based on a two-degree-of-freedom test of our priori-identified single primary intervention outcome measure (VO_2peak_) at two time points. We will use a contrast matrix to test both time points simultaneously, so no multiplicity adjustment will be needed. For all analyses, the Type I error rate will be set to α=0.05, and we will use a contrast matrix to conduct the F-test of our primary research hypothesis, which notably mimics a two-sided testing paradigm to even further protect against spurious results.

We anticipate the effect size (mean difference divided by the SD) to be half the effect of that observed in our 6-month trial[54] given the intensity of exercise will likely be lower than observed in a tightly controlled research protocol. Additionally, we anticipate a 20% decrease in this effect size at the 52-week endpoint compared to 12 week effect. We assume that the 12- and 52-week time point results will be positively correlated; thus, the probability of rejecting the null hypothesis of no group difference at both 12 weeks (Rej_l2 wks_) and 52 weeks (Rej_52 wks_) is Pr(Rej_l2 wks_ ∩ Rej_52 wks_) = Pr(Rej_l2 wks_|Rej_52 wks_) Pr (Rej_52 wks_)

We use this form of the joint distribution because we anticipate the effect size to be lower for the 52-week time point; hence our estimates of power are conservative. To estimate the joint probability of rejecting both we anticipate that if the effect is sustained (i.e., Pr [Rej_52 wks_], then the intervention group is highly likely to have increased VO_2peak_ at the 12-week time point. Thus, under the condition Rej_52 wks_, the conditional probability of Rej_l2 wks_, or Pr (Rej_l2 wks_|Rej_52 wks_) should be high (∼1.0). Therefore, Pr(Rej_l2 wks_|Rej_52 wks_) Pr (Rej_52 wks_) ≈ Pr (Rej_52 wks_). Using this, we can approximate the power based on data for a single time point.

In our prior study, we observed an effect size of 1.2 in the exercise intervention groups.[54] We conservatively assume the intervention effect will be approximately half that (0.6), and as indicated above we anticipate the 52-week time point to have a 20% decrease for the sustained effect, reducing the anticipated effect size to 0.48. With that our sample of 110 subjects per group will have over 88% power while allowing for 20% attrition (leaving approximately 88 subjects per group). With even smaller effect sizes (e.g., 0.45) this sample size will still have 84% power (nQuery Advisor® 7.0, 1995-2007).

### 2.9 Outcomes and Planned Statistical Analyses

This study aims to assess the implementation and scalability of the LEAP!Rx Program for clinicians and patients. Implementing the program requires both patient and clinician satisfaction. To asses the implementation and scalability of the LEAP!Rx Program for clinicians and patients. We will test the efficacy of an exercise and healthy lifestyle program using a unique referral method embedded in the electronic health record (EHR) as compared to the standard of care.

Secondarilty, we hypothesize that the LEAP! Rx Program will increase and maintain cardiorespiratory fitness (peak oxygen consumption [VO_2peak_]) at both 12 weeks (after the initiation phase) and 52 weeks (after the maintenance phase; co-primary outcome measure). VO_2peak_ is a vital health outcome linked to mortality and age-related diseases. It can be viewed as a physiological “assay” to prove that our intended health behavior change (increased physical activity as measured by accelerometry and survey) induces meaningful physiological benefits. We also expect increased moderate-to-vigorous physical activity (measured by accelerometry and survey) to be the mechanism responsible for VO_2peak_ benefits.

Finally, we are also interested in the effects of the LEAP! Rx Program on secondary outcome measures of chronic disease risk factors, including insulin resistance, body composition, and lipids. We hypothesize that LEAP! Rx Program will improve an individual’s metabolic profile with measurable benefits in specific secondary outcome measures of HOMA2, fat mass, lean mass, and cholesterol (total, LDL, HDL). Additional details on statistical analysis are available in Supplemental Material 2.

## 3.0 Conclusion

The ultimate goal of the LEAP! Rx study aims to create a scalable and cost-effective program for clinicians and their patients that reduces the risk of chronic disease by inducing lifestyle behavior change. The LEAP! Rx program begins with and supports the clinician-patient relationship with the intent to provide clinicians with a new tool composed of 1) a prescription, 2) a framework for delivering the prescription, and 3) metrics to assess effects on behavior. Recruitment of participants occurs through clinicians and is facilitated by an electronic prescription through the EHR that seamlessly integrates with the clinician’s workflow.

We use several methods to support continued behavior maintenance in real-world settings (i.e., mobile health monitoring for real-time feedback, community-based centers, and regular follow-up) to overcome barriers to sustained behavior change. The approach of the LEAP!Rx study is designed to realign common resources to allow scaling the program to essentially any staffed fitness center. We draw on our YMCA’s experience in scaling other interventions (Diabetes Prevention Program) and technology that is soon be ubiquitous (i.e. smartphones or wearable computing). If effective in this rigorous clinical trial, we will deliver an innovative program nationally to improve population health.

## Data Availability

This is a protocol manuscript. No human subjects data is reported.

## Abbreviations

LEAP! Rx Program: Lifestyle Empowerment for Alzheimer’s Prevention
EHR: Electronic Health Record
m-Health: mobile-health
AD: Alzheimer’s Disease
KU ADRC: University of Kansas Alzheimer’s Disease Research Center
IPAQ: International Physical Activity Questionnaire
6MWT: 6-Minute Walk Test
DEXA: dual-energy X-ray absorptiometry
HR-QOL: Health-related quality of life

## NCT Registration

NCT03253341

## Supplemental Material 1

### 2.4.1 LEAP! Rx Coach

Each fitness center will designate a LEAP!Rx Coach. The program design is based on the Behavioral Choice Theory. [80] LEAP! Rx Coaches will use motivational interviewing to assess readiness for change, motivations, goals, and barriers to physical activity. LEAP! Rx Coaches will have the following responsibilities:

- Function as the primary point of contact for study staff and participants
- Set individualized goals, monitor m-Health physical activity levels, review progress, and provide counseling on exercise, nutrition, and body weight.
- Lead exercise training sessions for supervision and training in resistance and aerobic exercise
- Attend monthly study meetings with area LEAP! Rx Coaches and study staff

### 2.4.2 LEAP! Coach Training and Qualifications

LEAP!Rx Coaches will have a personal trainer or fitness instructor certification from a nationally recognized organization. In addition, LEAP! Rx Coaches are required to complete the 4 phase training of ACE Integrated Fitness Training Model certification. This ACE certification prepares Coaches to deliver custom, individualized exercise programs with goals ranging from simple function to health, fitness, and elite performance.

### 2.4.3 Training Fidelity Plan

To enhance the fidelity of the delivery of the LEAP! Rx Program across fitness centers and to prepare LEAP! Rx Coaches for the broad health issues of an older population:

- LEAP! Rx Coaches will undergo a training program conducted by study personnel. The basis of this training will include a manual of procedures (MOP) modified from the YMCA’s recently developed “Coaching Connection” manual. This includes training on theories of behavior change and techniques for motivational interviewing. In the event of staff turnover, new LEAP! Rx Coaches are trained individually by study personnel.
- LEAP! Rx Coaches are trained in the exercise protocol and receive a manual developed over our several previous YMCA-based exercise trials. The manual will outline proper procedures for safe aerobic and resistance training progression and instruct on data collection and adverse event reporting.
- Ongoing training will be supported by close communication between LEAP! Rx Coaches and study staff. The LEAP! Rx study coordinator will lead monthly meetings bringing together the LEAP! Rx Coaches from the YMCA centers will discuss the program with study staff and troubleshoot any problems.
- We will intermittently (monthly) monitor coaches for protocol compliance and professionalism. Booster training sessions will be developed and employed as needed.

### 2.4.4 Exercise Training Program

The LEAP! Rx exercise program consists of an intensive 12-week Empowerment Phase followed by a 40-week Lifestyle Phase (See Supplemental Table 1.). The overall goal of the training is to meet current recommendations of 150 minutes a week of aerobic exercise (over 3 to 5 days a week) and 2 days a week of resistance exercise.[81, 82] To facilitate independence and self-reliance, personal training supervision will decrease in frequency throughout the 12-week Empowerment phase followed by monthly sessions during the Lifestyle phase. Participants are expected to increasingly exercise independently (home or at the YMCA) to meet the overall goal. The gradual introduction and progressively increasing volume and intensity of exercise are designed to build self-reliance.

LEAP! Rx Coaches will supervise in-person exercise training sessions. Coaches will also work with participants to set individualized goals and monitor progress toward those goals. One-on-one coaching will occur as part of the exercise training during the Empowerment phase and monthly afterward. Coaches will work to identify barriers and provide guidance for achieving the participant’s physical activity, fitness, nutrition, and other LEAP! Rx goals. LEAP! Rx Coaches will encourage participation in monthly educational sessions and group exercise opportunities.

In the event of an emergent situation, individuals may need to exercise from home with limited monitoring. Study staff will provide publicly or privately available exercise videos online and instructive handouts and will make as many check-in calls as possible. Videos are scientifically vetted or come from highly reputable sources (e.g. AHA or ACSM) and will attempt to support the existing protocol as much as possible. Online platforms are used to communicate with participants using a variety of means, including email and phone, as well as standard computer and phone apps that could provide a platform for face-to-face communication. These apps may include FaceTime, Skype, Zoom, or Microsoft Teams or similar apps. If these are used, we will do so on consultation with investigators who have ongoing telehealth research and apply their standard procedures for privacy. Handouts will encourage exercises that are consistent with public health recommendations and publications. Check-in call frequency will vary based on the situation, but we will target a biweekly call.

### 2.4.5 Empowerment Phase

The Empowerment phase comprises 12 weeks of supervised and unsupervised exercise training sessions. Aerobic exercise duration will be gradually increased over six weeks to a goal of 150 minutes per week. Exercise will be primarily supervised in the Empowerment phase and follow our protocol used in NIH-funded trials [19, 83] to successfully introduce and increase the volume of exercise in older adults. Participants will perform 60 total minutes of aerobic exercise in the first week and increase that volume by about 18 minutes per week to achieve 150 minutes per week by 6 weeks. Coaches will supervise during this titration period two times per week from Weeks 1-6 and one time per week in Weeks 8, 10 & 12. By Week 6, participants will walk, cycle, or elliptical for 150 minutes a week over 4-5 days at a level deemed “somewhat hard” or 3 to 5 (out of 10) on the modified Borg Rating of Perceived Exertion scale (RPE). [84] Use of an exertion rating as a proxy for a target heart rate zone decreases complexity, allows greater individualization in the context of bradycardic drugs and is a validated method of exercise prescription. Participants are taught how to rate their exertion levels based on their heart rate during supervised sessions.

Participants will also be encouraged to perform resistance exercises twice a week. Resistance exercise will target the major muscle groups (e.g., pectorals, rhomboids, and latissimus dorsi, biceps, triceps, quadriceps, hamstrings, calves). During Weeks 1-6, LEAP! Rx Coaches will work with participants to identify exercises and machines that are comfortable for the participant. Resistance will be set using the RPE to a resistance that is “somewhat hard” (RPE 3-5) to lift ten times. RPE is a valid method for gauging resistance training intensity like aerobic training.[85] Coaches will counsel participants on modifying exercises and resistance as participants increase strength.

Participants are encouraged to attend group exercise classes to meet the 150 minutes per week of aerobic exercise and two days per week of resistance training. Silver Sneakers classes are promoted as a good choice for participants.

### 2.4.6 Lifestyle Phase

The Lifestyle phase will consist of 40 weeks of once-monthly (every four weeks) exercise training sessions. Contact with the LEAP! Rx Coach will encourage adherence to the exercise program, assure the progression of exercise routines, and allow continual review of progress toward goals and monitoring of the m-Health data. These one-on-one training sessions will include continued goal-setting sessions to review goals, progress towards goals, and discuss other lifestyle factors such as nutrition, body weight, smoking cessation, etc. The goal of the Lifestyle phase is to continue to safely challenge the participant whiRx le allowing greater independence in the timing and location of exercise.

### 2.4.7 Mobile-Health (m-Health) Monitoring

The LEAP! Rx Program will leverage emerging m-Health technology to objectively measure physical activity and provide this data to the participant (self-monitoring), LEAP! Coaches, and ultimately the referring clinician (i.e., physical activity as a vital sign). Each participant will be given an m-Health monitoring device worn on the wrist (Garmin Vivofit2, Garmin, Olathe, KS). Devices store up to 30 days of data and have a battery life of 1 year. Data from these devices are collected through smartphone, computer or wireless access points (Garmin Vivohub2) that are installed at each of the YMCA sites and the participants smartphone (if available). Simply walking by a wireless access point will convey this data to our centralized database; thus, personal smartphones and computers are not required. LEAP! Rx Coaches will access an online customized dashboard (MyInertia) that provides them with access to their participants’ data. LEAP! Rx Coaches will either print a monthly hard copy or electronically view the physical activity report for their participants so that smartphones and computers are not required to access their data. At the end of study participation, LEAP! Rx Coaches will train the participants, whenever possible, to access their data through a device (i.e., smartphone or tablet) or on their home computer so they can continue using the devices.

### 2.4.8 Group Exercise Sessions

Each YMCA has a variety of group exercise opportunities available to YMCA members. Participants are encouraged to attend group exercise classes to augment their individualized, supervised exercise training sessions or home-based independent exercise. Group exercise opportunities are categorized into Beginner, Intermediate, and Advanced classes so that participants can appropriately progress their exercise volume and intensity. Beginner-level courses are encouraged during the initiation phase, Intermediate during weeks 12 to 24, and Advanced during weeks 25 to 52. LEAP! Rx Coaches will assist in identifying classes that will effectively fulfill aerobic and resistance exercise goals.

### 2.4.9 Smart Aging Educational Curriculum

The Smart Aging Educational Curriculum will be delivered in person at the KU Clinical Research Center by KU ADRC study staff with an alternate option for viewing online. The overall goal of the curriculum will be to initiate and sustain behavior change and provide a framework for lifestyle enhancement. Although each session will include some didactic information, the sessions will primarily focus on group discussion and goal-setting. The Smart Aging Curriculum has been developed and deployed at the KU ADRC. We will use a local media company to create and translate the existing KU ADRC programming into consumer-friendly and professionally packaged materials. To support the delivery of the program, YMCA staff will receive training materials. Sessions will include online videos guided completion of a workbook with a LEAP! Rx Coach that: 1) asks participants to reflect on their habits indicated by monitoring and changes with age, 2) sets realistic and measurable goals, describes expectations for change in behaviors and barriers to making change, 3) evaluates perceived benefits of increasing activity, reinforces strategies for rewarding behavior change and enjoyable activities to pursue, ratings of confidence in the ability to make changes, 4) tips for ways to change behavior (generic and targeted to individual habits/barriers). The educational and goal-setting piece is based on Social Cognitive Theory and Behavioral Choice Theory concepts. [80]

**Supplemental Table 1.** LEAP! Rx Program Events.

| Initiation Phase |  |  |  | Maintenance Phase |
| --- | --- | --- | --- | --- |
|  | Weeks<br>1-4 | Weeks<br>5-8 | Weeks<br>8, 10, 12 | Weeks 13 to 52 |
| **Supervised Exercise Training** | 3 per<br>week | 2 per<br>week | 1 per<br>week | Every 4 weeks (10 sessions) |
| **Group Exercise** | Weekly | Weekly | Weekly | Weekly (40 sessions) |
| **Smart Aging Curriculum Class** | Monthly | Monthly | Monthly | Every 4 weeks (9 sessions) |

**Table 2.** Study Assessments.

|  |  |  | Visit 1 |  | Visit 2 |  | Visit 3 |
| --- | --- | --- | --- | --- | --- | --- | --- |
|  | Referral |  |  | 12-week<br>Empowerment<br>Phase |  | 36-week<br>Lifestyle<br>Phase |  |
|  | Week - 4 |  | Baseline |  | Week 12 |  | Week 52 |
| **Study Events** |  |  |  |  |  |  |  |
| Chart Review -- Meds, PMH | X |  |  |  |  |  |  |
| Phone Screen (Health Hx, TAPA) | X |  |  |  |  |  |  |
| Consent |  |  | X |  |  |  |  |
| Peak VO2 |  |  | X |  | X |  | X |
| DEXA / Height, weight |  |  | X |  | X |  | X |
| PA assessment - accelerometry |  |  | X |  | X |  | X |
| Fasting Blood Draw |  |  | X |  | X |  | X |
| NIH Emotion, Cognitive Toolbox, PROMIS |  |  | X |  | X |  | X |
| NCI Diet History Questionnaire 2, Veggie Meter |  |  | X |  | X |  | X |
| Randomization |  |  | X |  |  |  |  |

**Table 3:**
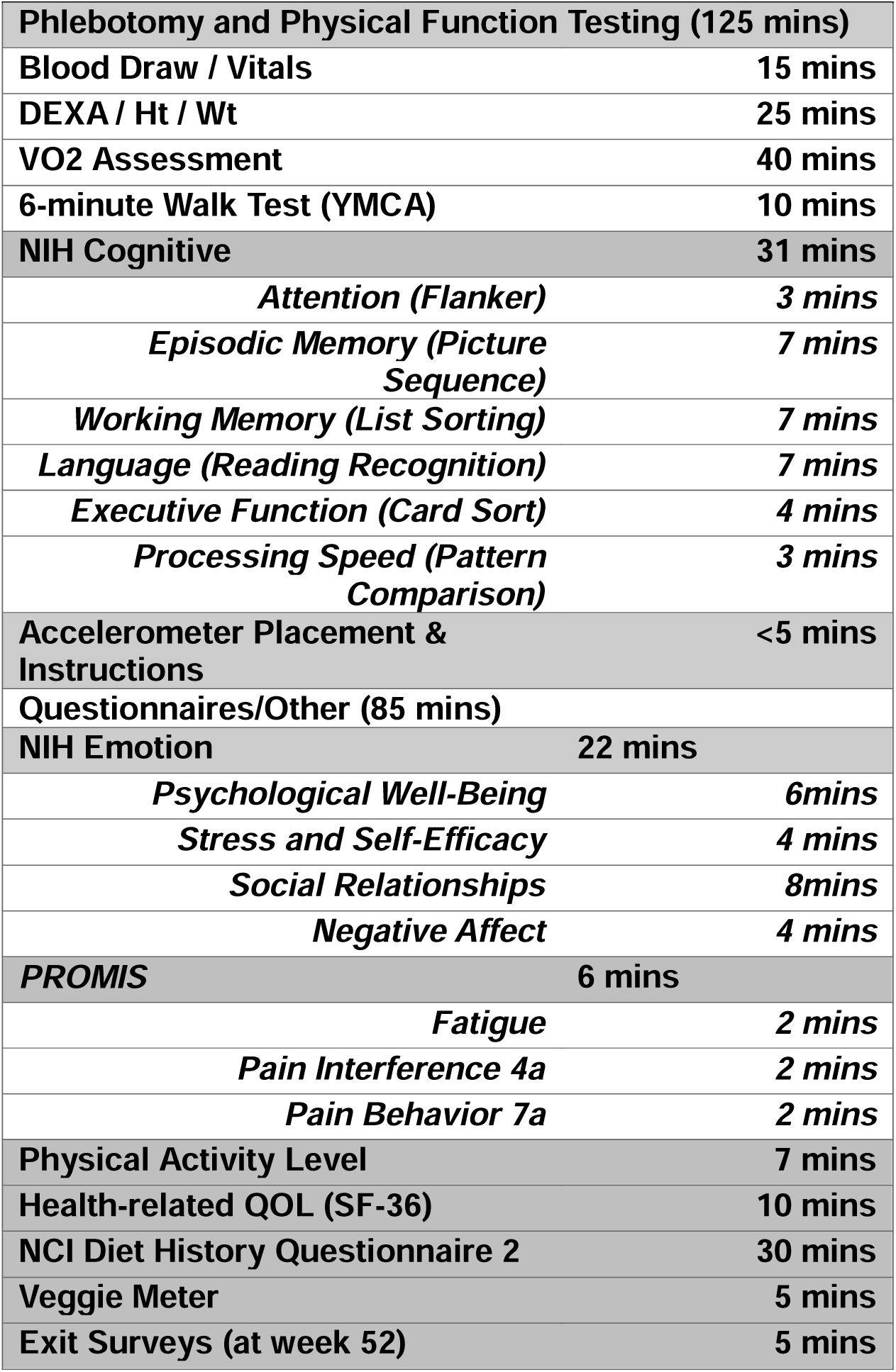
Outcome Measures and their Duration.

| **Table 3: Outcome Measures and their Duration** |  |
| --- | --- |
| **Phlebotomy and Physical Function Testing (125 mins)** |  |
| Blood Draw / Vitals | 15 mins |
| DEXA / Ht / Wt | 25 mins |
| VO2 Assessment | 40 mins |
| 6-minute Walk Test (YMCA) | 10 mins |
| **NIH Cognitive** | **31 mins** |
| *Attention (Flanker)* | *3 mins* |
| *Episodic Memory (Picture Sequence)* | *7 mins* |
| *Working Memory (List Sorting)* | *7 mins* |
| *Language (Reading Recognition)* | *7 mins* |
| *Executive Function (Card Sort)* | *4 mins* |
| *Processing Speed (Pattern Comparison)* | *3 mins* |
| Accelerometer Placement & Instructions | <5 mins |
| **Questionnaires/Other (85 mins)** |  |
| **NIH Emotion** | **22 mins** |
| *Psychological Well-Being* | *6mins* |
| *Stress and Self-Efficacy* | *4 mins* |
| *Social Relationships* | *8mins* |
| *Negative Affect* | *4 mins* |
| **PROMIS** | **6 mins** |
| *Fatigue* | *2 mins* |
| *Pain Interference 4a* | *2 mins* |
| *Pain Behavior 7a* | *2 mins* |
| Physical Activity Level | 7 mins |
| Health-related QOL (SF-36) | 10 mins |
| NCI Diet History Questionnaire 2 | 30 mins |
| Veggie Meter | 5 mins |
| Exit Surveys (at week 52) | 5 mins |

## Supplemental Material 2

1. To asses the implementation and scalability of the LEAP!Rx Program for clinicians and patients. We will test the efficacy of an exercise and healthy lifestyle program using a unique referral method embedded in the electronic health record (EHR) as compared to the standard of care.

Test the effect of the LEAP! Rx Program on outcome measures of chronic disease risk factors, including insulin resistance, body composition, and lipids. We hypothesize that LEAP! Rx Program will improve an individual’s metabolic profile with measurable benefits in specific secondary outcome measures of HOMA2, fat mass, lean mass, and cholesterol (total, LDL, HDL).

### 2.9.1 Aim 1

Study Aim 1 is to asses the implementation and scalability of the LEAP!Rx Program for clinicians and patients. We will test the efficacy of an exercise and healthy lifestyle program using a unique referral method embedded in the electronic health record (EHR) as compared to the standard of care.

Descriptive statistics will be calculated for referral status, randomization group, retention, and demographics. Statistical tests will be conducted to compare differences in referral status, retention, and demographics and include chi-squared tests for categorical variables and t-tests for continuous variables.

Patient-centered outcomes will also involve generalized linear mixed models (GLMMs) at baseline, 12-, and 52-week time points. For continuous measures, the LMMs (a special case of the GLMMs) will suffice. In contrast, other more discrete-measured variables will require different model assumptions (such as the multinomial distribution with an ordinal logit function, similar to that for the clinician measures), such as with Likert-type measures. Model assessments will include residual analyses (to assess mean-to-variance relationships and whether they are in keeping with the exponential family distribution selected for the particular GLMMs) and observed versus expected comparisons. We will utilize the two degree-of-freedom testing paradigms previously described to assess sustained benefit simultaneously and better control type I errors

### 2.9.2 Aim 2

The goal of Aim 2 is to determine the effect of the LEAP! Rx Program on cardiorespiratory fitness. We hypothesize that the LEAP! Rx Program will increase and maintain cardiorespiratory fitness (peak oxygen consumption [VO_2peak_]) at both 12 weeks (after the initiation phase) and 52 weeks (after the maintenance phase). Descriptive statistics will be generated for all measures. Continuous measures will include the mean, standard deviation, percentiles, and range; categorical measures will include frequencies and relative frequencies. Bivariate analyses comparing the two treatment groups will consist of visual inspection (e.g., box plots for continuous measures) and statistical tests such as the two-sample t-test, Pearson’s chi-square test, or analogous nonparametric measures as indicated based on assessments of underlying assumptions (e.g., residual analysis for t-tests, expected cell counts for Pearson’s chi-square tests).

The primary study measure, VO_2 peak_, will have repeated measures over time (baseline, week 12, and week 52). Thus, we will use linear mixed models (LMMs) for analysis. Estimated linear contrasts will assess change (baseline to 12 weeks **and** baseline to 52 weeks) in VO_2_ peak comparing active vs. control groups, providing a randomized, intent-to-treat comparison. Our primary research hypothesis (Aim 1) involves a two degree-of-freedom test. To conclude with our research hypothesis of a benefit at 12 weeks sustained at 52 weeks, a contrast matrix will simultaneously test in a single statistical test that VO_2peak_ increases over baseline at 12 weeks and 52 weeks.

We will use a type I error rate of 0.05 for this simultaneous two-degree-of-freedom comparison of the primary measure (F-test). Tests of secondary measures, hypotheses, and per-protocol analyses (based on _≥_80% adherence) will follow a similar LMM approach. Further, we will collect detailed medical history and medications (using well-developed methods) to account for these potential confounding variables. Specific attention will be paid to beta blockers and steroids and cardiovascular co-morbidities as potential modifiers, given their known influence on our primary cardio-metabolic outcomes. As these factors can be initiated or changed during the trial, we will incorporate them as time-varying covariates within our proposed LMM approach. Our initial strategy for these changes will be to add an indicator (time-varying) for whether their medication or co-morbidity was initiated, increased, or decreased. Since we have identified and controlled the type I error for our two primary assessments (via the single, simultaneous two-degree-of-freedom F-test), no further multiplicity adjustments for our measures specified a priori as secondary will be used. We will fully disclose the measures tested and their a priori classification as primary versus secondary when reporting our findings in the literature to control the type I error.

Analyses will be performed using statistical software such as SAS and R. LMMs will be assessed by residual analysis (predicted vs. residual plots, q-q plots, etc.), visual inspection of residuals, and comparison of estimated standard deviations across groups (for homogeneity of variance). If indicated, our model assessment will use alternative strategies (transformation, bootstrapping, etc.).

### 2.9.3 Aim 3

The goal of Aim 3 is to test the effect of the LEAP! Rx Program on chronic disease risk factors, including insulin resistance (HOMA2), body composition, and lipids (total cholesterol, LDL / HDL). We hypothesize the LEAP! Rx Program will positively affect an individual’s metabolic profile with measurable benefits in insulin resistance, reduced fat mass / increased lean mass, and lipid status.

The secondary study measures for aim 2 include HOMA2, lean mass, fat mass, total cholesterol, HDL, and LDL at baseline, 12-, and 52-week time points; thus, similar analysis methods as in aim 1 will be used (LMMs). Assessments for these measures will examine change from baseline to 12 weeks and whether any benefits will be sustained at 52 weeks via a single statistical test. These two degree-of-freedom tests will better control the operating characteristics study-wide. For each measure, we will assess the corresponding LMM by residual analysis (predicted vs. residual plots, q-q plots, etc.), visually inspect residuals, and compare estimated standard deviations across groups (for homogeneity of variance). As indicated, these assessments will use alternative strategies (transformation, bootstrapping, etc.).

